# Population-based assessment of neurodevelopmental and mental health diagnoses among pediatric patients with Turner Syndrome: A PEDSnet study

**DOI:** 10.1101/2025.06.17.25329800

**Authors:** Christa Hutaff-Lee, Morgan Jolliffe, Karli S Swenson, Holly Wakeman, Deanna Swain, Anna Furniss, Natalie Nokoff, Jen Hansen-Moore, Chijioke Ikomi, Vaneeta Bamba, Rachel E. Lean, Skyler Leonard, Shanlee M Davis

## Abstract

Individuals with Turner syndrome (TS) are known to be at increased risk for neurodevelopmental disorders (NDD) and mental health (MH) conditions, but data from large, population-based pediatric samples remain limited. We examined the prevalence of NDD and MH diagnoses among youth with TS (N = 2,145) compared to matched female controls (N = 8,580) across six U.S. pediatric health systems. Odds ratios (OR) and 95% confidence intervals (CI) were calculated using generalized estimating equations. Youth with TS had significantly higher odds of an NDD diagnosis (24.2% vs. 11.9%; OR 2.37, 95% CI 2.11–2.67), particularly for speech-language, motor, learning, and attentional disorders. Increased odds were also observed for autism spectrum disorder (ASD) and intellectual developmental disorder (IDD), though these remained relatively uncommon. In contrast, MH diagnoses, such as anxiety and mood disorders, were not more prevalent in TS compared to controls (17.3% vs. 18.5%; OR 0.92, 95% CI 0.81–1.05). These findings support the need for proactive neurodevelopmental screening in TS and raise important questions about the recognition and documentation of MH conditions in this population. Additional research is warranted to understand whether MH symptoms are underdiagnosed in youth with TS or emerge later in development.

## INTRODUCTION

Turner syndrome (TS) is a genetic disorder that occurs in 1:2000-2500 females due to a partial or complete absence of the second sex chromosome.^1,2^ Karyotypes vary, with approximately half of individuals presenting with non-mosaic 45,X.^3^ Phenotypic heterogeneity in TS is influenced by the specific karyotype as well as other genetic and environmental factors. Classically, TS is associated with short stature, congenital and acquired cardiac conditions^4.5^, primary ovarian insufficiency, and sensorineural hearing loss.^6^ Standard treatments often include growth hormone and estrogen replacement therapy. In addition to these physical manifestations, neurodevelopmental disorders (NDD) and mental health (MH) conditions are also common among individuals with TS.

Existing research characterizing the neurocognitive profile in TS reveals an increased likelihood of developmental and cognitive challenges. Developmental delays across fine motor, gross motor, and language domains are commonly reported in TS.^7^ A recent multi-site study found that 40.9% of 631 children with TS had documented developmental concerns in their medical records.^8^ Most individuals with TS have overall intellectual abilities within the average range, although significant discrepancies between domains are often present.^9^ Verbal reasoning is often a relative strength, while visual-spatial deficits are frequently noted – previously characterized as the verbal IQ-performance IQ (VIQ-PIQ) split.^10^ Attention-related challenges are also prevalent, with increased rates of Attention-Deficit/Hyperactivity Disorder (ADHD) reported, with studies estimating that 14-25% of individuals with TS have ADHD.^8,11,12^ However, there is high variability among studies, with a large population-based study from Sweden finding only 1.6% of individuals with TS had ADHD compared to 1.2% of controls.^12^ Additionally, specific learning disabilities, particularly in mathematics, affect 43-55% of individuals with TS.^13,14^ Social communication deficits, especially understanding facial expressions or nonliteral/figurative language (e.g., sarcasm, idioms), are also frequently reported.^15^ In one large population study, individuals with TS were four times more likely to have an autism spectrum disorder (ASD; 2% vs <1% in the general population).^12^ Another study found that 3% of individuals with TS met the diagnostic criteria for ASD, approximately twice the incidence observed in the general female population.^12^ Some neurodevelopmental outcomes may be directly or indirectly impacted by medical comorbidities. For example, children with congenital heart disease requiring surgery or prolonged hospitalization are at increased risk for NDDs due to a variety of risk factors that can impact the developing brain, including the potential for chronic hypoxia.^16^ Similarly, hearing impairment can place a child at increased risk for delays in speech-language and literacy development.^17^ Despite well-documented NDD comorbidities in TS, population-based data on the relative odds of these conditions compared to peers remain limited.

MH conditions concerning mood/affective problems are also commonly associated with TS. Reported prevalence rates vary by population and study design, ranging from 22% to 70%, with depression and anxiety being most frequently observed.^15,18,19^ Most studies suggest the prevalence of MH conditions is higher in TS than typical female peers; however, disparities diminish when compared to females with short stature^20^ or premature ovarian insufficiency.^12,21^ Thus, as with NDDs, MH concerns may be more closely related to the medical phenotype of TS than to intrinsic genetic risk. The impact of MH disorders across development in TS remains unclear. A recent systematic review noted increased risk for depression in adolescents and adults with TS, but not in children.^22^ A recent chart review spanning three large pediatric centers found that 20.4% of youth with TS had documented MH concerns; however, the study lacked a direct comparison group.^8^ In contrast, Avdic et al (2021) found only 7% of individuals with TS were diagnosed with an anxiety disorder compared to 8% of the control group.^12^ Taken together, these findings highlight the need for further research into the association between MH disorders and TS, particularly in pediatric populations.

To date, much of the literature on NDD and MH outcomes in TS is derived from small, clinic-based samples. The current study aimed to expand on those findings using data from a large, diverse U.S. pediatric cohort, with diagnostic and treatment information reflective of U.S.-based clinical practice. We hypothesized that youth with TS would have increased odds of receiving NDD and MH diagnoses compared to matched peers.

## MATERIALS AND METHODS

Data Source: PEDSnet is a pediatric clinical research network facilitating collaborative, patient-centered pediatric research in the U.S. Six children’s health systems – Children’s Hospital Colorado, Children’s Hospital of Philadelphia, Nemours Children’s Health System, Nationwide Children’s Hospital, St. Louis Children’s Hospital, and Seattle Children’s Hospital participated in this project, collectively representing over 6 million children. A limited dataset including individual-level data for patient demographics, clinical encounter types, diagnoses (from billing codes and/or problem list), and prescription medications from electronic health records (EHR) was obtained from the PEDSnet Data Coordinating Center. This project was approved by the PEDSnet Research Committee and was determined by the local Institutional Review Boards to be non-human subject research, as research activities were limited to receipt and analysis of deidentified data.

Analytic Sample: Patients within participating PEDSnet institutions with a diagnosis code mapping to TS (see Supplemental Table 1 for codes previously validated) and at least one outpatient visit from 2009-2019 were included as TS Cases (n=2,145) as per our prior TS computable phenotype analysis.^23^ A random sample of 113,725 females with at least one outpatient visit and no diagnosis of TS or any other genetic condition were used as a pool of controls. Because demographic variables were dissimilar between TS cases and controls, we proceeded with propensity score matching. Propensity scores were generated via multivariable logistic regression based on year of birth, age at last encounter, PEDSnet site, race, ethnicity, health insurance payer status (public/private/none), and duration in the PEDSnet database (time between first and last encounter). Missing data were handled by the widely accepted approach of including missing as separate category for the variable. Each TS case was matched to four controls on the logit of the propensity score using a greedy match algorithm and a caliper of width 0.1. Covariate balance was evaluated using standardized mean differences, with values less than 0.2 indicative of acceptable balance.

**Table 1.** Cohort Demographics.

|  | TS (%)<br>N=2,145 | Controls (%)<br>N=8,580 | p-value |
| --- | --- | --- | --- |
| Site |  |  | 0.93 |
| Site 1 | 252 (11.7) | 1,043 (12.2) |  |
| Site 2 | 423 (19.7) | 1,612 (18.8) |  |
| Site 3 | 290 (13.5) | 1,135 (13.2) |  |
| Site 4 | 552 (25.7) | 2,230 (26.0) |  |
| Site 5 | 426 (19.9) | 1,745 (20.3) |  |
| Site 6 | 202 (9.4) | 815 (9.5) |  |
| Race |  |  | >0.99 |
| White | 1,458 (68.0) | 5,823 (67.9) |  |
| Black | 179 (8.3) | 696 (8.1) |  |
| Asian | 54 (2.5) | 215 (2.5) |  |
| Other | 284 (13.2) | 1,158 (13.5) |  |
| Unknown | 170 (7.9) | 688 (8.0) |  |
| Ethnicity |  |  | 0.68 |
| Hispanic | 311 (14.5) | 1,240 (14.5) |  |
| non-Hispanic | 1,704 (79.4) | 6,775 (79.0) |  |
| Unknown | 130 (6.1) | 565 (6.6) |  |
| Health insurance status |  |  | 0.04 |
| Public | 820 (38.2) | 3,317 (38.7) |  |
| Private | 1,119 (52.2) | 4,433 (51.7) |  |
| Other | 156 (7.3) | 543 (6.3) |  |
| Unknown | 50 (2.3) | 287 (3.3) |  |
| Age at first visit (years) | 4.4 [0.3, 10.2] | 4.3 [0.7, 9.9] | 0.05 |
| Age at last visit (years) | 14.3 [8.5, 18.5] | 14.6 [10.0, 17.2] | 0.08 |
| Duration in PEDSnet (years) | 7.1 [2.8, 11.5] | 7.3 [2.2, 12.2] | 0.48 |
| Total # of outpatient visits | 17.0 [8.0, 38.0] | 7.0 [3.0, 20.0] | <0.01 |
| Age at first TS diagnosis (years) | 8.0 [2.0, 13.0] | n/a | n/a |
| Age at last TS diagnosis (years) | 13.0 [7.0, 17.0] | n/a | n/a |
| # of encounters with a TS diagnosis | 10.0 [4.0, 22.0] | n/a | n/a |
| *Data are presented as N (%) or median [IQR]* |  |  |  |

Outcomes: The PEDSnet Common Data Model directly maps to SNOMED-CT concept codes, an international system for clinical terminology. Investigators refined composites to encompass the diagnoses of interest (Supplemental Table 1). The NDD Composite included diagnoses related to developmental delays, autism spectrum disorders, learning disorders, and behavioral disorders. The MH Composite included all conditions falling under the SNOMED parent code “Mental Disorder” except for the concept “developmental mental disorder,” “developmental delay in feeding,” “feeding disorder of infancy,” and “non-organic infant feeding disturbance”. Additional composites were also developed to assess related outcomes of interest including sleep disorders, failure to thrive, and self-harm. In addition to diagnoses, RxNorm codes were used to query prescriptions for any stimulant medications, selective serotonin uptake inhibitors (SSRIs) and antipsychotic medications. All outcomes were treated as binary variables; if there was no documentation of the condition within the record it was determined to not be present.

Statistical analyses: Descriptive statistics (frequencies and percent, means and standard deviations and/or median with interquartile ranges) were used to summarize sample demographics and outcomes. Generalized estimating equations (GEE) were used to compare TS cases to controls, and the odds ratio (OR) and 95% confidence intervals were reported. In accordance with PEDSnet policy, cells with fewer than 11 individuals were not reported. In addition to the GEE models, logistic regression models including covariates of growth hormone prescription (yes/no) and estrogen prescription (yes/no) were conducted for the TS cases. Given the large dataset and multiple comparisons, conservative significance threshold of p<0.0025 was applied. All statistical analyses were conducted in SAS version 9.4 (SAS Institute Inc., Cary, NC).

## RESULTS

There were 2,145 individuals within PEDSnet with a diagnosis of TS. The median age at first TS diagnosis was 8 years; 42.3% had a documented prescription for growth hormone, and 32.2% had a prescription for estrogen. Demographics were similar between TS and their matched controls, although patients with TS had more total outpatient visits (Table 1). Significantly more patients with TS had encounters with a behavioral health provider (37.6% for TS vs 24.2% for controls, p<0.0001).

### NDD Outcomes

Nearly one quarter (24.2%) of individuals with TS had a diagnosis encompassed by the NDD Composite, representing a significantly higher odds compared to controls (OR 2.4 [95% CI 2.11 - 2.67]). The TS group had a higher prevalence of all diagnoses falling under the NDD Composite, with the exception of disruptive behavior disorders (Table 2). Longer duration in the PEDSnet system and younger age at diagnosis were associated with higher odds of diagnoses within the NDD Composite; however inclusion of these variables in the model did not change the results. Within the TS group, treatment with growth hormone was positively associated with diagnoses within the NDD composite, including learning disabilities and ADHD. In addition, use of estrogen was positively associated with a learning disorder diagnosis (Supplemental Table 2). Although individuals with TS were at a higher risk for being diagnosed with ADHD, prescriptions for stimulant medication were not significantly different across the two groups (5.3% vs. 4.1%).

**Table 2.** NDD and MH Diagnoses.

|  | TS<br>N=2,145 | Controls<br>N=8,580 | OR (95% CI) | p-value |
| --- | --- | --- | --- | --- |
| NDD Composite | 520 (24.2) | 1,020 (11.9) | 2.37 (2.11, 2.67) | **<0.0001** |
| Development delay composite | 188 (8.8) | 231 (2.7) | 3.47 (2.86, 4.22) | **<0.0001** |
| Motor delay disorder composite | 245 (11.4) | 511 (6.0) | 2.04 (1.73, 2.39) | **<0.0001** |
| Feeding delay disorder composite | 138 (6.4) | 149 (1.7) | 3.89 (3.07, 4.94) | **<0.0001** |
| Speech and language disorder composite | 241 (11.2) | 408 (4.8) | 2.54 (2.15, 2.99) | **<0.0001** |
| Learning disorder composite | 95 (4.4) | 129 (1.5) | 3.04 (2.34, 3.95) | **<0.0001** |
| Disruptive behavior disorder | 101 (4.7) | 345 (4.0) | 1.18 (0.94, 1.48) | 0.15 |
| Attention deficit hyperactivity disorder | 190 (8.9) | 453 (5.3) | 1.74 (1.46, 2.08) | **<0.0001** |
| Intellectual developmental disorder | 51 (2.4) | 118 (1.4) | 1.75 (1.26, 2.41) | **0.0007** |
| Autism spectrum disorder | 72 (3.4) | 122 (1.4) | 2.41 (1.79, 3.23) | **<0.0001** |
| **MH Composite** | **371 (17.3)** | **1,586 (18.5)** | **0.92 (0.81, 1.05)** | **0.21** |
| Adjustment disorder | 32 (1.5) | 162 (1.9) | 0.79 (0.54, 1.15) | 0.22 |
| Anxiety disorder | 138 (6.4) | 680 (7.9) | 0.80 (0.66, 0.96) | 0.02 |
| Obsessive-compulsive disorder | 13 (0.6) | 57 (0.7) | 0.91 (0.51, 1.64) | 0.76 |
| Eating disorder | 32 (1.5) | 143 (1.7) | 0.89 (0.61, 1.32) | 0.57 |
| Mood disorder | 78 (3.6) | 647 (7.5) | 0.46 (0.36, 0.59) | **<0.0001** |
| Depressive disorder | 67 (3.1) | 578 (6.7) | 0.45 (0.34, 0.58) | **<0.0001** |
| Tic disorder | 17 (0.8) | 67 (0.8) | 1.02 (0.59, 1.74) | 0.96 |
| **Other related diagnoses** |  |  |  |  |
| Sleep disorder | 279 (13.0) | 698 (8.1) | 1.69 (1.46, 1.95) | **<0.0001** |
| Self-harm composite | 14 (0.7) | 192 (2.2) | 0.29 (0.17, 0.50) | **<0.0001** |
| **Encounters and Prescriptions** |  |  |  |  |
| Behavioral Health Encounter | 806 (37.6) | 2,072 (24.2) | 1.89 (1.71 - 2.09) | **<0.0001** |
| SSRI Prescription | 87 (4.1) | 455 (5.3) | 0.75 (0.60 - 0.95) | 0.018 |
| Antipsychotic Prescription | 42 (2.0) | 259 (3.0) | 0.64 (0.46 - 0.89) | 0.007 |
| Stimulant Prescription | 113 (5.3) | 353 (4.1) | 1.30 (1.04-1.61) | 0.029 |

*Data are presented as N (%). TS = Turner syndrome; NDD = neurodevelopmental disorder; MH = mental health; SSRI = selective serotonin reuptake inhibitor.*

### MH Outcomes

In contrast to the NDD Composite, youth with TS had similar prevalance of MH diagnosis than matched controls (17.3% vs 18.5%, p=0.21). Longer study duration and older age at last follow up appointment were associated with higher odds of diagnoses within the MH composite; however adding these variables to the model did not change results. Compared with controls, patients with TS had a *lower* prevalence of both anxiety (6.4% vs 7.9%, OR 0.80 [0.66, 0.96] and mood disorders (3.6% vs 7.5%, OR 0.46 [0.36, 0.59]). Similarly, self-harm diagnoses were less common among youth with TS (OR 0.29 [95% CI 0.17 – 0.50]) compared to controls. Congruent with diagnostic findings, there was no increased use of SSRIs (4.1% vs 5.3%) or antipsychotic medications (2.0% vs 3.0%) in patients with TS compared to controls. Within the TS cohort, neither growth hormone nor estrogen prescriptions were associated with any MH diagnoses (Supplemental Table 2). Although not strictly classified as a MH condition, sleep disorders were more prevalent among youth with TS (13.0% vs 8.1%, OR 1.69 [95% CI 1.46 - 1.95]) and were positively associated with growth hormone treatment.

**Figure 1.**
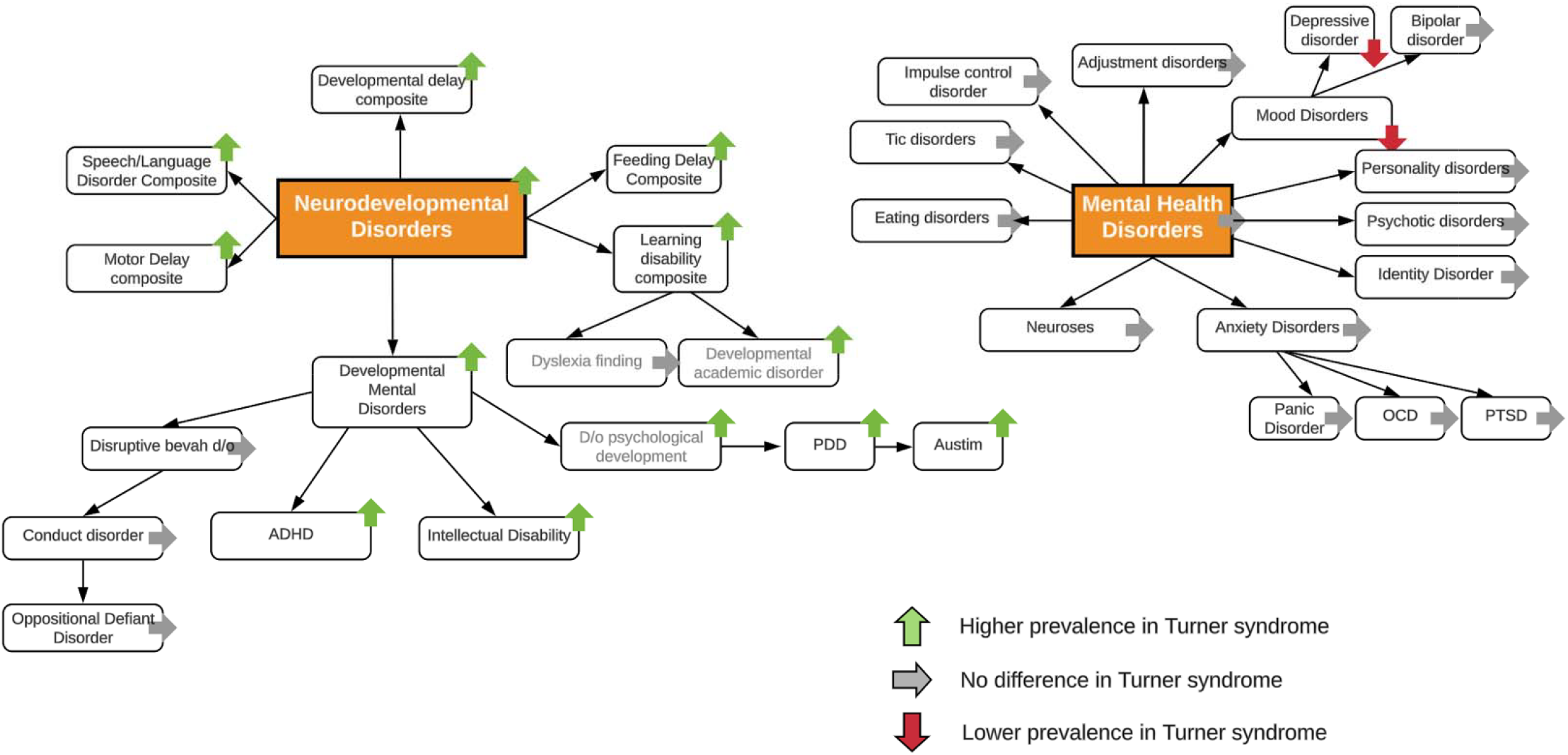
Neurodevelopmental disorder (NDD) diagnoses are diffusely increased in youth with Turner syndrome ( compared to age- and sex-matched peers, however mental health (MH) diagnoses are not.

## DISCUSSION

This large, multicenter U.S. study of pediatric patients with TS demonstrates that youth with TS are at significantly increased risk for NDDs compared with matched peers, but not for MH diagnoses. One in four individuals with TS had an NDD diagnosis, with particularly elevated odds for speech-language, motor, learning, and attention disorders. In contrast, the prevalence of MH diagnoses, such as anxiety and mood disorders, was comparable or even lower than in matched controls. These findings suggest that while NDDs are more consistently recognized and diagnosed in TS, MH dianoses may not be fully captured in EHR data. If youth with TS are at higher risk for MH conditions, as clinical experience would suggest, there is a need for improved screening and diagnostic practices to ensure these concerns are identified and addressed in clinical care.

Our findings regarding NDDs are consistent with previous literature describing increased neurodevelopmental risk in TS. Diagnoses of learning disabilities and ADHD were significantly more common among individuals with TS in this study, supporting prior studies that document these difficulties as part of the TS neurocognitive phenotype.^12^ Intellectual developmental disorder was more prevalent in TS compared to controls, although overall rates remained relatively low, consistent with the broader consensus that intellectual functioning in TS is generally within the average range. ^9,14^ ASD was similarly more prevalent in TS, although rates remain low. The observed ADHD prevalence (9%) was higher than in the Swedish registry study (1.6%)^12^ but lower than in smaller U.S. clinic-based cohorts reporting rates as high as 25%.^11^ This suggests that while increased attentional difficulties are recognized in some individuals with TS, they may still be underdiagnosed, particularly if symptoms are attributed to the TS phenotype rather than conceptualized as a separate disorder.

Despite the elevated rate of ADHD diagnoses, stimulant prescriptions were not more common in the TS group than in controls. This may reflect under-treatment or under-recognition of functionally impairing attention difficulties, a phenomenon observed in other pediatric populations with chronic health conditions. For example, research among cancer survivors has shown that many children experience attentional impairments without meeting formal diagnostic criteria for ADHD.^24^ The emerging use of “neurodevelopmental disorder associated with a medical condition” as a diagnostic label highlights the need for greater diagnostic nuance in medically complex populations. Our findings emphasize the importance of systematic screening and referral to neuropsychological services for youth with TS, as recommended by the TS Clinical Practice Guidelines ^25^, to ensure that cognitive and behavioral symptoms are appropriately evaluated and supported.

In contrast to expectations and clinical experience, we did not find an increased prevalence of MH diagnoses, such as anxiety or mood disorders, among individuals with TS. Rates of self-harm diagnoses and prescriptions for SSRIs and antipsychotics were also similar or lower than in controls. While this pattern was surprising, it mirrors findings from another large population-based study by Avdic et al. (2021), which also found no increased risk of anxiety or related disorders among individuals with TS compared to the general population.^12^ One potential explanation is that MH conditions are under-recognized or under-documented in pediatric medical settings. Many youth with TS are followed primarily by specialists such as endocrinologists or cardiologists, who may not routinely screen for emotional or behavioral concerns. A recent survey of pediatric endocrinologists reported low confidence in identifying and addressing MH concerns among patients with TS.^26^ In addition, behavioral health providers may bill for psychosocial concerns using Health and Behavior codes, which would not appear as psychiatric diagnoses in the EHR.^27^ It is also possible that the expression of MH symptoms in TS differs from typical presentations, complicating recognition using standard diagnostic frameworks. Prior studies have suggested that anxiety in TS may present with somatic or internalized features that are less likely to trigger formal diagnosis.^28^ Another important consideration is the nature of our control group, which was limited to pediatric patients seen at tertiary care institutions. Many of these children likely have chronic medical conditions, which are themselves associated with elevated MH risk. This may have attenuated differences between groups and contributed to the relatively high rate of MH diagnoses in controls. Indeed, both groups had MH diagnosis rates below the 25% prevalence estimated for adolescent anxiety in the U.S.,^29^ which may reflect underdiagnosis or limited documentation of MH conditions in subspecialty care.

In addition to group-level differences, we examined individual-level factors associated with diagnostic patterns in TS. Longer duration in the PEDSnet system was associated with higher odds of both NDD and MH diagnoses, which was also observed in a similar study among youth with differences of sex development suggesting that increased time in care provides more opportunities for symptom recognition and documentation.^30^ Age patterns also aligned with clinical expectations: younger age was associated with greater odds of NDD diagnoses, reflecting the early developmental onset of many of these conditions, while older age was associated with increased likelihood of MH diagnoses, which may emerge, or be more readily identified, during adolescence.^31^ We also found that treatment with growth hormone and estrogen was associated with higher odds of NDD diagnoses. These associations likely reflect phenotype severity rather than causal effects of treatment, as individuals requiring these therapies may represent a more medically complex subset of the TS population. Taken together, these findings highlight how developmental stage, medical complexity, and duration of clinical follow-up may influence the likelihood of receiving a documented NDD or MH diagnosis in TS.

## Limitations

Several limitations should be considered. First, the use of deidentified EHR data limited our ability to stratify results by karyotype or mosaicism status, which may impact neurodevelopmental risk. Although prior studies suggest strong diagnostic validity of TS in PEDSnet,^23^ we could not confirm specific genetic findings or differentiate more complex presentations (e.g., ring X chromosome). Second, reliance on billing codes may have resulted in misclassification or missed diagnoses, especially for conditions that are less consistently documented or diagnosed in specialty care. Third, care delivered outside of PEDSnet institutions, including community or school-based evaluations, was not captured, likely leading to underestimates of NDD and MH prevalence. We were also unable to control for important risk factors such as prematurity, anesthesia exposure, or medical complexity, which are known to affect neurodevelopmental outcomes.

Finally, PEDSnet policy on small cell sizes (<11 individuals) restricted reporting on rare conditions.

## Conclusion

In this large, population-based study of youth with Turner syndrome, we found significantly increased odds of NDD diagnoses, especially in the areas of speech-language, learning, motor, and attention, compared to matched controls. However, we did not observe increased rates of documented MH diagnoses, raising the possibility that these concerns are under-recognized or underreported in pediatric specialty care. These findings support the need for routine, proactive screening for both NDD and MH concerns in youth with TS. As research continues to clarify the neurocognitive and social-emotional phenotype associated with TS, it will be important to identify modifiable risk factors and develop targeted interventions to improve long-term outcomes.

**Supplemental Table 1.**
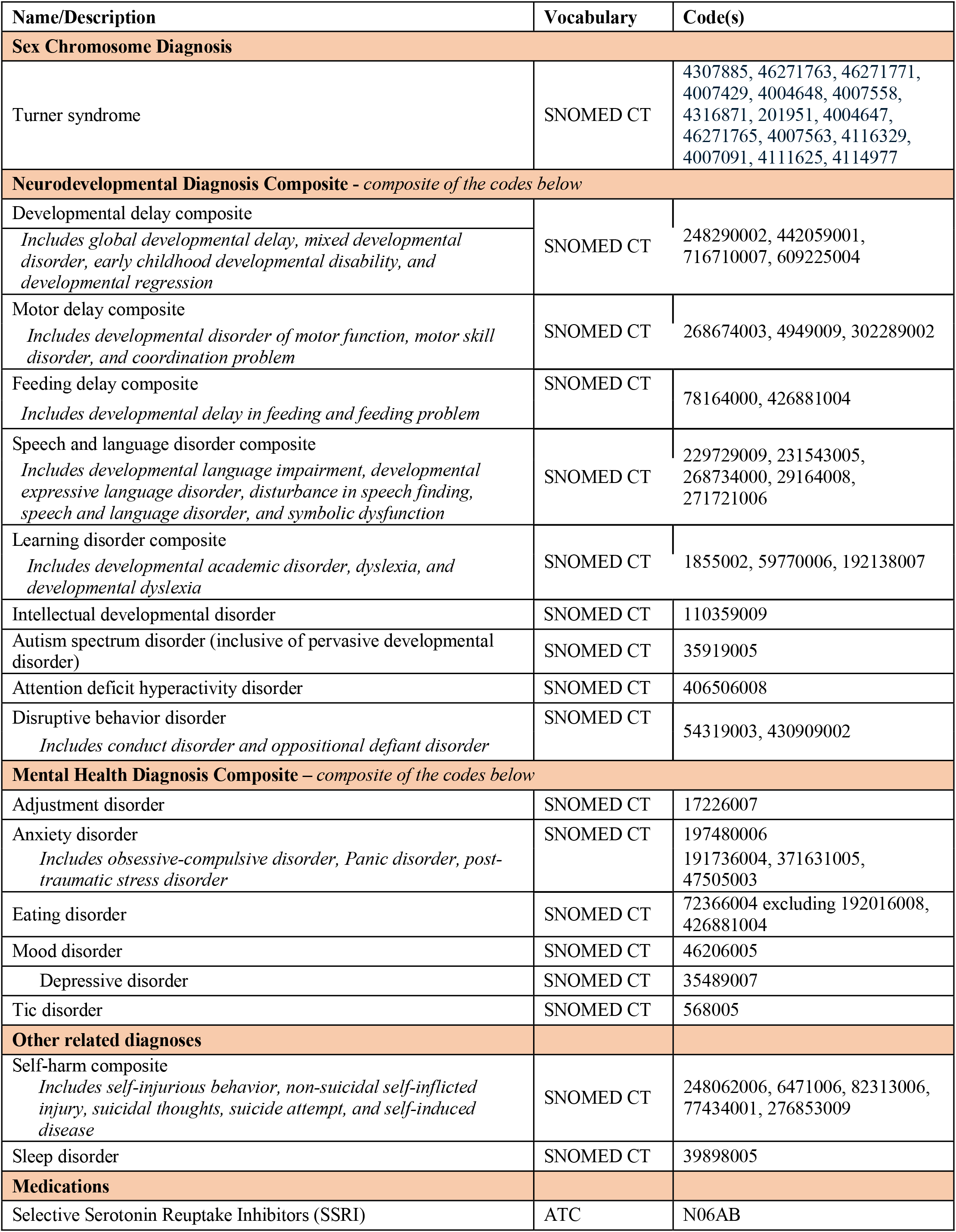
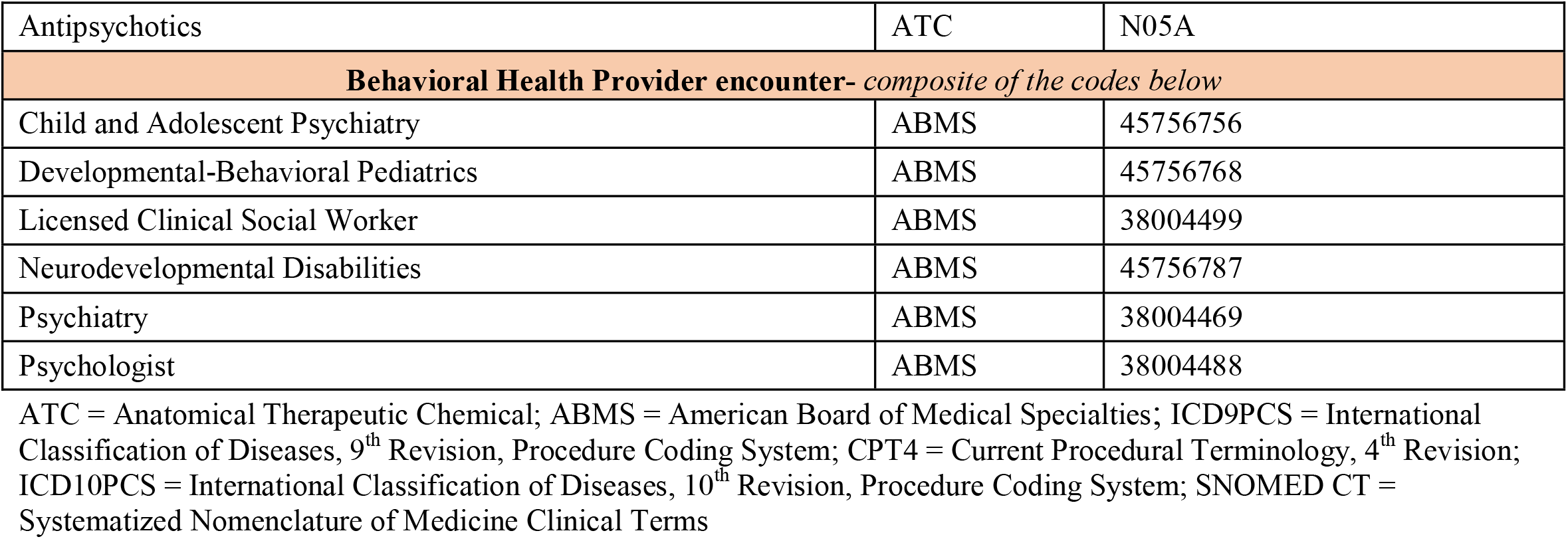
Definitions.

**Supplemental Table 2.** Associations between estrogen or growth hormone prescriptions and outcomes, TS only.

|  | Estrogen |  | Growth hormone |  |
| --- | --- | --- | --- | --- |
|  | OR (CI) | p value | OR (CI) | p value |
| **NDD Composite** | **0.87 (0.67, 1.12)** | **0.29** | **1.37 (1.11, 1.68)** | **0.004** |
| Development delay composite | 0.77 (0.51, 1.17) | 0.22 | 1.20 (0.86, 1.66) | 0.29 |
| Motor delay disorder composite | 0.95 (0.68, 1.32) | 0.77 | 1.42 (1.08, 1.87) | 0.01 |
| Feeding delay disorder composite | 0.70 (0.34, 1.45) | 0.34 | 0.79 (0.49, 1.28) | 0.34 |
| Speech and language disorder composite | 0.71 (0.48, 1.06) | 0.09 | 1.42 (1.05, 1.91) | 0.02 |
| Learning disorder composite | 1.97 (1.20, 3.22) | 0.007 | 3.68 (2.31, 5.86) | **<0.0001** |
| Disruptive behavior disorder | 1.03 (0.65, 1.61) | 0.91 | 1.50 (0.99, 2.26) | 0.06 |
| Attention deficit hyperactivity disorder | 0.98 (0.69, 1.39) | 0.90 | 1.71 (1.26, 2.33) | **0.0006** |
| Intellectual developmental disorder | 0.52 (0.27, 1.01) | 0.05 | 0.47 (0.25, 0.88) | 0.02 |
| Autism spectrum disorder | 1.03 (0.51, 2.06) | 0.93 | 0.86 (0.48, 1.56) | 0.62 |
| **MH Composite** | **0.93 (0.72, 1.21)** | **0.59** | **1.18 (0.94, 1.49)** | **0.15** |
| Adjustment disorder | 1.40 (0.60, 3.28) | 0.43 | 2.12 (1.03, 4.34) | 0.04 |
| Anxiety disorder | 0.75 (0.50, 1.12) | 0.16 | 0.99 (0.69, 1.41) | 0.94 |
| Eating disorder | 1.05 (0.49, 2.29) | 0.89 | 0.98 (0.48, 1.99) | 0.95 |
| Mood disorder | 0.83 (0.51, 1.35) | 0.45 | 0.58 (0.36, 0.94) | **0.03** |
| Depressive disorder | 0.87 (0.52, 1.46) | 0.60 | 0.61 (0.36, 1.03) | 0.07 |
| Tic disorder | 0.74 (0.25, 2.15) | 0.58 | 1.11 (0.43, 2.89) | 0.83 |
| **Other related diagnoses** |  |  |  |  |
| Sleep disorder | 1.08 (0.77, 1.50) | 0.66 | 2.01 (1.53, 2.64) | **<0.0001** |
| Failure to thrive | 1.30 (0.88, 1.93) | 0.19 | 2.08 (1.52, 2.85) | **<0.0001** |

## Data Availability

Data has limited availability, but can be accessed through formal request and approval through PEDSnet.

## Acknowledgements

We would like to thank the PEDSnet Data Coordinating Center for their support in the data acquisition and all PEDSnet site contributors.

